# Family Research of Microbes Linked to Respiratory Infections (FAMILY Micro) observational study: Assessing the use of minimally invasive self-sampling methodologies at home for long-term monitoring of the oral, nasal and hand microbiota of adults and children within UK families

**DOI:** 10.1101/2023.04.19.23288393

**Authors:** E. Nikolaou, E.L. German, A. Howard, H.M. Nabwera, A. Matope, R. Robinson, F. Shiham, K. Liatsikos, C. McNamara, S. Kattera, K. Carter, C.M. Parry, J.M. Read, S.J. Allen, B.C. Urban, D.B. Hawcutt, H. Hill, A.M. Collins, D.M. Ferreira

## Abstract

**Background:** Monitoring the presence of commensal and pathogenic microorganisms in the human oral, nasal and hand niches as determinants for respiratory tract infections is of critical global relevance as was evident during the COVID-19 pandemic. However, community-based surveillance is difficult because current sampling methods are not optimal for a wide age range of participants, particularly young children. We designed a platform of minimally invasive self-sampling at home and assessed its use for longitudinal monitoring of the oral, nasal and hand microbiota of adults and children within families.

**Methods:** Healthy families with two adults and up to three children, living in and near Liverpool, United Kingdom, self-collected saliva, nasal lining fluid using synthetic absorptive matrices and hand swabs at home every two weeks for six months. Questionnaires were used to collect demographic and epidemiological data and assess feasibility and acceptability. At the end of the study, participants were invited to take part in an interview.

**Results:** Thirty-three families completed the study. Sample collection using our approach was acceptable to 25/33 (76%) families, as sampling was fast (76%), easy (76%) and painless (60%). Saliva and hand sampling was acceptable to all participants of any age, whereas nasal sampling was accepted mostly by adults and children older than 5 years.

**Conclusion:** Multi-niche self-sampling at home can be used by adults and children for longitudinal microbial surveillance, providing key data for monitoring respiratory infections.

## Introduction

Respiratory tract infections (RTI) affect the sinuses, throat, airways, and lungs [1]. Lower RTIs are the leading cause of death in children under five worldwide [2, 3] and the third leading cause of death in all ages in the United Kingdom (UK) [4, 5]. A quarter of the population of England and Wales visit their GP because of an RTI each year [6]. RTIs are responsible for 60% of all antibiotic prescriptions in primary care, which constitutes a significant cost to the national health system [7] and contributes to antimicrobial resistance [8]. Common causes of RTIs are the bacterium *Streptococcus pneumoniae* [3, 4] (pneumococcus) and a plethora of viruses which include respiratory syncytial virus for bronchiolitis in children globally [9], coronavirus SARS-CoV-2 for adult pneumonia, and seasonal influenza. RTI occurrence changes with host characteristics, geographical region, environmental factors, antibiotic use, and lifestyle habits [10-13].

Healthy individuals carry commensal bacteria but can also asymptomatically carry pathogenic bacteria in their upper respiratory tract (URT). Together, the microbiota, is likely to influence respiratory health [14] and is constantly shared through person to person contact especially within a household [15]. Interactions with viruses, antibiotics and other factors can alter bacterial composition and are likely to influence transmission of bacterial pathogens between household members. Although the presence and abundance of pathogenic bacteria are strongly associated with susceptibility and severity of RTIs in children [11-13], it is unclear why some people get mild or severe disease, while others remain asymptomatic when infected with the same pathogen.

Research so far has focused on the relationship between the respiratory microbiome, including viruses, and RTI occurrence and severity by comparing healthy to RTI-infected individuals (single measurement case-control studies). Typically, these involve mothers and children up to 5yrs [10-13]. This is because large community-based surveillance studies involving repeated sampling of individuals across a wide age range are difficult to conduct due to the nature of the currently recommended URT sampling methods. To detect pneumococcal carriage in the URT, a pre-requisite for disease occurrence, the World Health Organisation (WHO) recommends the use of nasopharyngeal swabs (NPS) in children and both NPS and oropharyngeal swabs in adults [16]. However, sampling, especially in children, is challenging as nasopharyngeal swabs can cause significant discomfort and pain. Saliva and nasal lining fluid using synthetic absorptive matrices (SAM) are less invasive and have been used successfully for paediatric sampling for both bacterial [17] and viral [18] detection.

Our group established self-sampling at home using saliva and SAM for studying pneumococcal carriage after experimental exposure in adults [19]. We have also investigated the use of SAM for the detection of pneumococcal carriage in hospitalised children aged 1-5 years under anaesthesia [20], showing that SAM has equal sensitivity to the current gold standard NPS [21]. Moreover, we have demonstrated that hands can be vehicles for transmission of pneumococcus and lead to acquisition of nasopharyngeal carriage [22]. This study employs our previously established methodologies of saliva, nasal fluid and hand sampling and adapts them for longitudinal monitoring of the oral, nasal and hand microbiota in families. Our monitoring platform also included questionnaires for capturing risk factors for RTI occurrence and transmission. Here, we present the study design and its acceptability to study participants. We seek to establish saliva, nasal lining fluid using SAM and hand sampling for monitoring microbial presence and transmission in the community with a scope to identify an “at risk” microbiome for RTIs.

## Material and Methods

### Study design and subjects

The FAMILY Micro study (ISRCTN 52814289) was a collaboration between the Liverpool School of Tropical Medicine and the local paediatric hospital, Alder Hey Children’s Hospital, that was ethically approved by the North West - Greater Manchester West Research Ethics Committee (20/NW/0304). Healthy families comprising of two adults aged 18-60 years and between one to three children aged 28 days – 18 years living in or close to Liverpool, UK were enrolled between October 2020 and August 2021. In our previous home sampling study in adults, 61/63 (97%) participants accepted and complied successfully with self-testing at home [19]. A sample size of 125 participants was therefore required to detect a 97% compliance rate with a 95% confidence interval and an error margin of 3%. Anticipating a dropout rate of approximately 20%, we adjusted the sample size target to a maximum of 160 participants.

The study design was adapted to adhere to COVID-19 restriction measures. Recruitment was conducted via email to staff in local hospitals and universities and parents in local schools and nurseries, flyers at clinics in Alder Hey Hospital, social media, and word of mouth. Consent and training appointments took place virtually over Microsoft Teams. First, a member of the clinical team assessed participants’ eligibility (Supplementary Table 1). Eligible participants were then trained in study procedures by a member of the research team who described sample collection by sample type and explained how to complete the questionnaires. A bag with sample kits and a laminated sheet with sample collection kit contents (Supplementary Material 1), an information sheet on sample collection (Supplementary Material 2), a laminated step-by step instructions sheet, a dated collection schedule (Supplementary Material 3) and a complete set of questionnaires were provided to participants after the consent appointment. Participants aged 16 years and above provided written informed consent prior to study commencement, as a photo or scanned email attachment before sending in the post if necessary. Children aged 11 to 15 years were asked to sign an assent form and for younger children, parental consent was covered by the parent’s consent form and verbal assent during the initial consent appointment as confirmed by the study doctors. Communication between the study team and participants was maintained throughout the study via the WhatsApp platform, including reminders of sampling dates, arranging times for sample pick-ups, and answering any queries.

### Sampling procedures

Parents and older children (5 years or older) self-collected saliva, nasal lining fluid and hand swabs at home once every two weeks for six months (Figure 1). Parents helped younger children (<5 years) to collect samples. Each sample kit was clearly labelled with study week number and distinguished between family members by participant ID (Parent A, Parent B, Child A, Child B or Child C) and a colour-coded label. Tubes with bacteria preservative media were kept in domestic fridges (4°C) until use. Family members were asked to collect samples on the same day as per their collection schedule or if that wasn’t possible, one day before or after the scheduled date. Participants were asked to send a WhatsApp picture of the samples to the study’s dedicated, password-protected phone to confirm collection. Samples were stored in domestic freezers (−20°C) until pick-up and transport to the research laboratory at the Liverpool School of Tropical Medicine. Pick-ups were arranged half-way through and at the end of the study with samples transported on cold packs. Participants were also given the option to request more frequent pick-ups as required.

**Figure 1:**
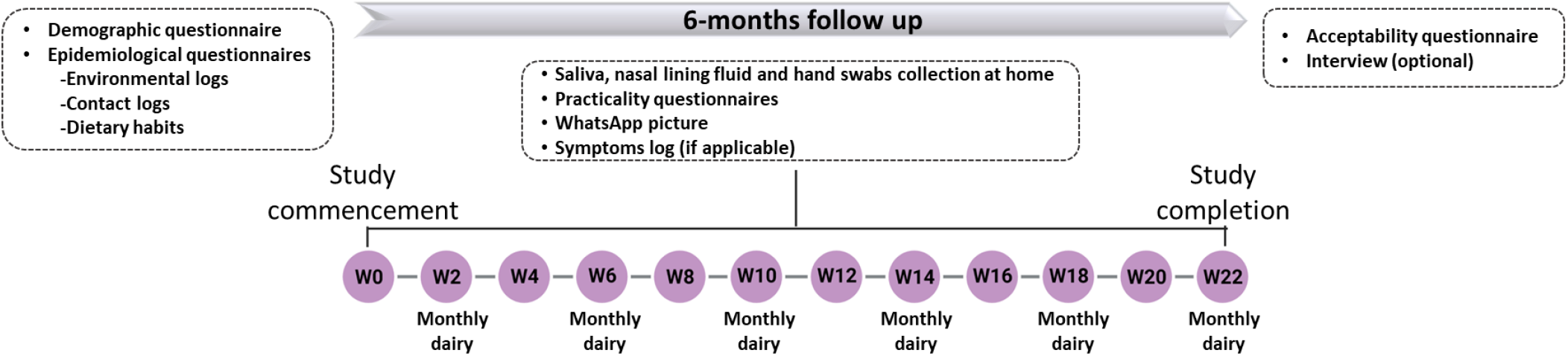
Timeline of the FAMILY Micro study.

#### Saliva collection

Participants were asked to spit into a 50mL centrifuge tube (Appleton Woods, UK) to collect approximately 1ml saliva at least 30 minutes after last drinking, eating, or brushing their teeth. Saliva was preserved in 1mL STGG (Skimmed milk – Tryptone – Glucose – Glycerine) 10% glycerol media [23], poured into the tubes after collection. For children <2 years that were likely unable to spit, chewable paediatric absorbent swabs (Salimetrics, USA) were used for saliva collection by holding them in their mouth for 2 minutes. Swabs were stored in accompanying centrifuge tubes (Sarstedt, Germany) pre-filled with STGG. Children aged 2-5 years were given the option between the two methods.

#### Nasal lining fluid collection

Participants were asked to hold the SAM strip (Hunt Development LTD, UK) inside one nostril for up to 2 minutes. The diameter of the strip used differed according to participants’ age (adults 7mm and children 4.5mm) as per manufacturer instructions. Infants (<1yr) were given an extra option for a smaller strip of 3mm diameter.

#### Hand swabs collection

Participants were asked to swab their unwashed dominant hand, using a sterile cotton swab (MWE, UK) after soaking it with sterile saline (Laboratoires Gilbert, France). Swabs were stored in accompanying tubes pre-filled with STGG.

### Quantitative and qualitative data collection

#### Questionnaires (quantitative)

Participants completed questionnaires throughout the study (Figure 1). Demographic and epidemiological (environmental log, contact log, dietary habits) data were collected at the beginning of the study. For assessing the feasibility of the sample collection methods, participants completed a questionnaire after every sampling time point to monitor each participant’s level of discomfort and pain and time taken to collect each sample type. For monitoring each family member’s health, participants were asked to complete a symptom log for each episode of illness. The family’s routines, such as means of transport and time spent at work, school, or home, were also recorded in monthly dairies. At the end of the study, participants completed an exit questionnaire on the acceptability of sample collection methods, indicating the overall opinion of the family, that of individual participants, and the key reasons for it (Supplementary Material 4).

#### Interview (qualitative)

At the end of the study, adult participants were invited for an optional interview to share their experiences (Figure 1). Interviews were performed via Microsoft Teams and recorded using the record meeting function. Recordings were stored in a secure study file on the research team’s shared drive and were deleted post-transcription. Results from interviews were used to assess the feasibility of our approach and are reported elsewhere [24].

### Data analysis

Baseline demographics were analysed using summary statistics. Continuous variables were summarised using the number of observations, mean, median, standard deviation, and minimum and maximum values, and for categorical (nominal) variables, the number and percentage of subjects was used. Data from children were discussed overall and in five age groups: infants (28 days – 1 year), toddlers (1-2 years), pre-schoolers (3-5 years), school-aged children (6-12 years) and adolescents (13-17 years).

The primary endpoints of the study were participants’ compliance and acceptability of the sample collection methods. To assess participant compliance, returned samples were counted and expressed as a percentage (%) of the total number of expected samples per sample type for each participant, based on the number of weeks their family took part in the study. Average compliance was calculated for each age group. Acceptability of home sampling was assessed based on the exit questionnaire. Sampling acceptability was assessed overall and per sample type. Overall acceptability was expressed as the % of families or participants who accepted home sampling out of the total number of families or participants, with 95% binomial confidence intervals. The reasons for acceptability were reported as % of families who indicated each reason. Fourteen families scored their top three reasons in order from most (with a score of 1) to least (with a score of 3) important. Here, we also present the % of these families who gave number 1 score per reason. We also calculated the participant acceptability per individual sample type using comments written on the exit questionnaire. Sample type acceptability was expressed as % of the total number of participants accepted each sample type.

## Results

### Recruitment approach summary

Eighty-four families were approached or expressed interest in participating in the FAMILY Micro study and were sent the Patient Information Leaflet (Figure 2). Of these, twenty-eight (33%) either refused or never replied, while a further fifteen (18%) were deemed ineligible. One family was recruited but never returned any samples or documents to the study team. Therefore, forty families (48%), representing a total of 157 participants, were monitored in the FAMILY Micro study (Figure 2). The study was conducted while COVID-19 restrictions were changing in UK. Twenty-five participating families (62%) were recruited through word of mouth by the study team and by existing participants. Communication emails to staff in local hospitals and universities (12 families, 30%) and to parents in local schools and nurseries (3 families, 8%) were also a successful recruitment strategy and these remained open despite COVID-19 restrictions. Recruitment via social media or flyers at Alder Hey Hospital clinics was unsuccessful.

**Figure 2:**
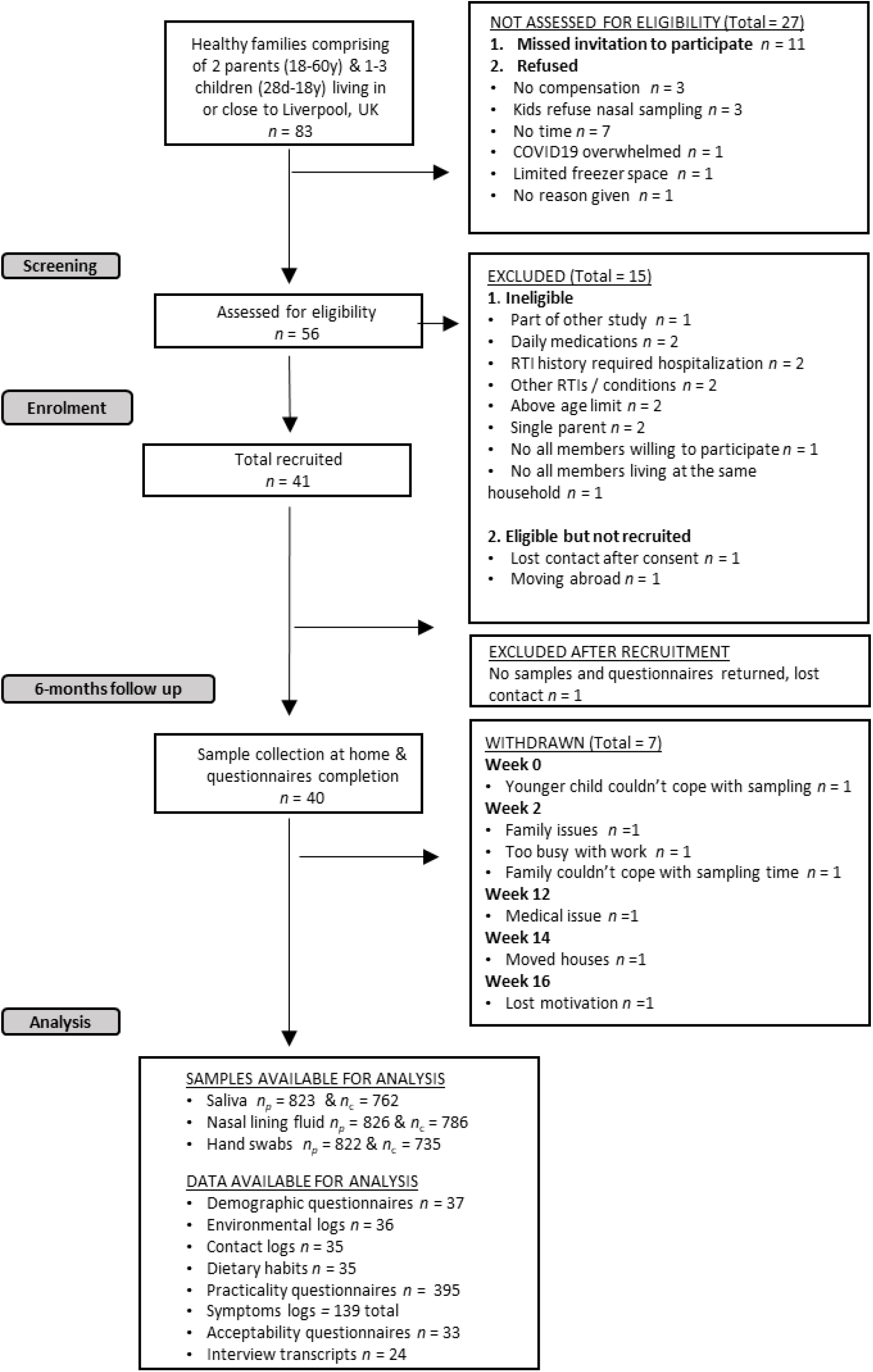
STROBE diagram for FAMILY Micro study. Number of families n, number of parents n_p_ and number of children n_c_

### Households and participants demographics

Most of the households were comprised of four family members (83%), were based in the Liverpool City Region (78%) and had an income range of £50,000-100,000 (78%) (Table 1). The mean age of parents was 39.8 years (range of 31-54 years) with a male:female ratio of 1:1. Most parents had completed university-level education (80%) and a high proportion (46%) were employed in medical, research and life science professions, as expected from the recruitment strategy (Table 2). The mean age of children (N=77) was 6.5 years (range of 2 months – 17 years) including 9 infants (mean age = 5.0 months), 11 toddlers (mean age = 1.5 years), 21 pre-schoolers (mean age = 4.2 years), 21 school-aged children (mean age = 8.5 years) and 15 adolescents (mean age = 14.1 years). Male children were 55% with the distribution varying by age group from 37% of infants being male compared to 62% of adolescents (Table 3).

**Table 1:**
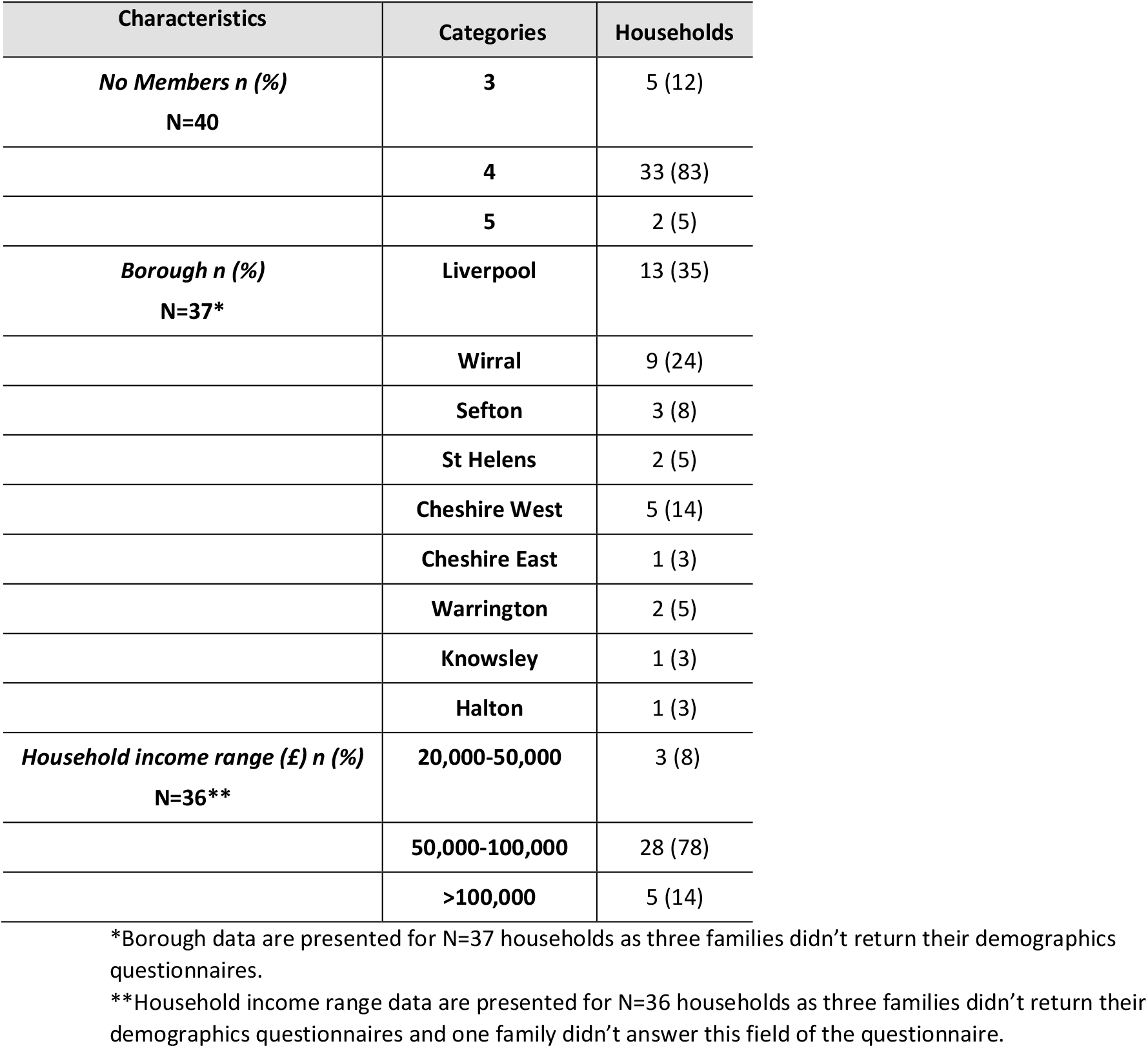
Summary of households’ demographics.

| Characteristics | Categories | Households |
| --- | --- | --- |
| **No Members n (%)**<br>**N=40** | **3** | 5 (12) |
|  | **4** | 33 (83) |
|  | **5** | 2 (5) |
| **Borough n (%)**<br>**N=37\*** | **Liverpool** | 13 (35) |
|  | **Wirral** | 9 (24) |
|  | **Sefton** | 3 (8) |
|  | **St Helens** | 2 (5) |
|  | **Cheshire West** | 5 (14) |
|  | **Cheshire East** | 1 (3) |
|  | **Warrington** | 2 (5) |
|  | **Knowsley** | 1 (3) |
|  | **Halton** | 1 (3) |
| **Household income range (£) n (%)**<br>**N=36\*\*** | **20,000-50,000** | 3 (8) |
|  | **50,000-100,000** | 28 (78) |
|  | **>100,000** | 5 (14) |

\*Borough data are presented for N=37 households as three families didn't return their demographics questionnaires.
\*\*Household income range data are presented for N=36 households as three families didn't return their demographics questionnaires and one family didn't answer this field of the questionnaire.

**Table 2:**
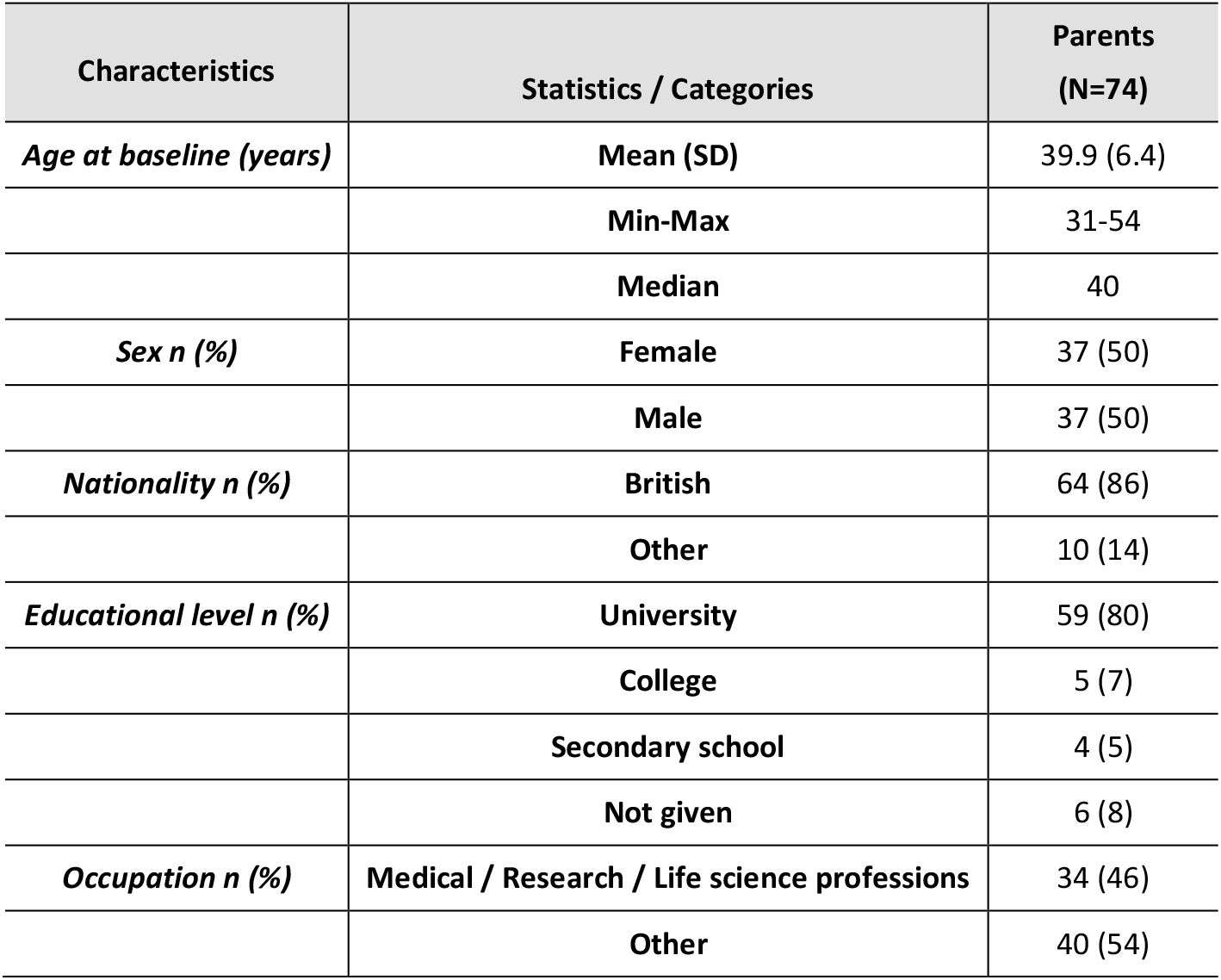
Summary of parents’ baseline demographics. Data is presented for N=74 parents as three families didn’t return their demographics questionnaires (six parents excluded).

**Table 3:** Summary of children’s baseline demographics. Data is presented per age group and in total.

|  |  | Children |  |  |  |  |  |
| --- | --- | --- | --- | --- | --- | --- | --- |
| Characteristics | Statistics / Categories | Infants (N=9) * | Toddlers (N=11) | Pre-schoolers (N=21) | School-aged (N=21) | Adolescents (N=15) | All (N=77) |
| *Age at baseline (years)* | **Mean (SD)** | 5.0m (2.5) | 1.5 (0.5) | 4.2 (0.8) | 8.5 (1.5) | 14.1 (1.4) | 6.5 (4.9) |
|  | **Min-Max** | 2m-9m | 1-2 | 3-5 | 6-12 | 13-17 | 2m-17 |
|  | **Median** | 4.0 | 2.0 | 4.0 | 9.0 | 14.0 | 5.0 |
| *Sex n (%) \*\** | **Female** | 5 (63) | 4 (40) | 10 (50) | 8 (40) | 5 (38) | 32 (45) |
|  | **Male** | 3 (37) | 6 (60) | 10 (50) | 12 (60) | 8 (62) | 39 (55) |
| *Nationality n (%) \*\** | **British** | 6 (75) | 10 (100) | 19 (95) | 20 (100) | 12 (92) | 67 (94) |
|  | **Other** | 2 (25) | 0 (0) | 1 (5) | 0 (0) | 1 (8) | 4 (6) |

\*Infants age in months
\*\*No sex or nationality were recorded for one infant, one toddler, one pre-schooler, one school-aged and two adolescents thus percentages are reported to the total number of records per child category rather than to the total number of children recruited per category (Infants N=8, Toddlers N=10, Pre-schoolers N=20, School-aged N=20, Adolescents N=13, All N=71).

### Methodology compliance

All 40 families and 157 participants were included in the analysis of compliance, with a total of 1612 hand, 1585 saliva and 1557 nasal lining fluid samples returned to the research laboratory. Compliance was over 80% for all sample types and age groups (Figure 3 A-C). Compliance was highest for hand swabs. In the youngest age groups (infants, toddlers, and pre-schoolers), compliance was lower for saliva and lowest for nasal lining fluid sampling (Figure 3C).

**Figure 3.**
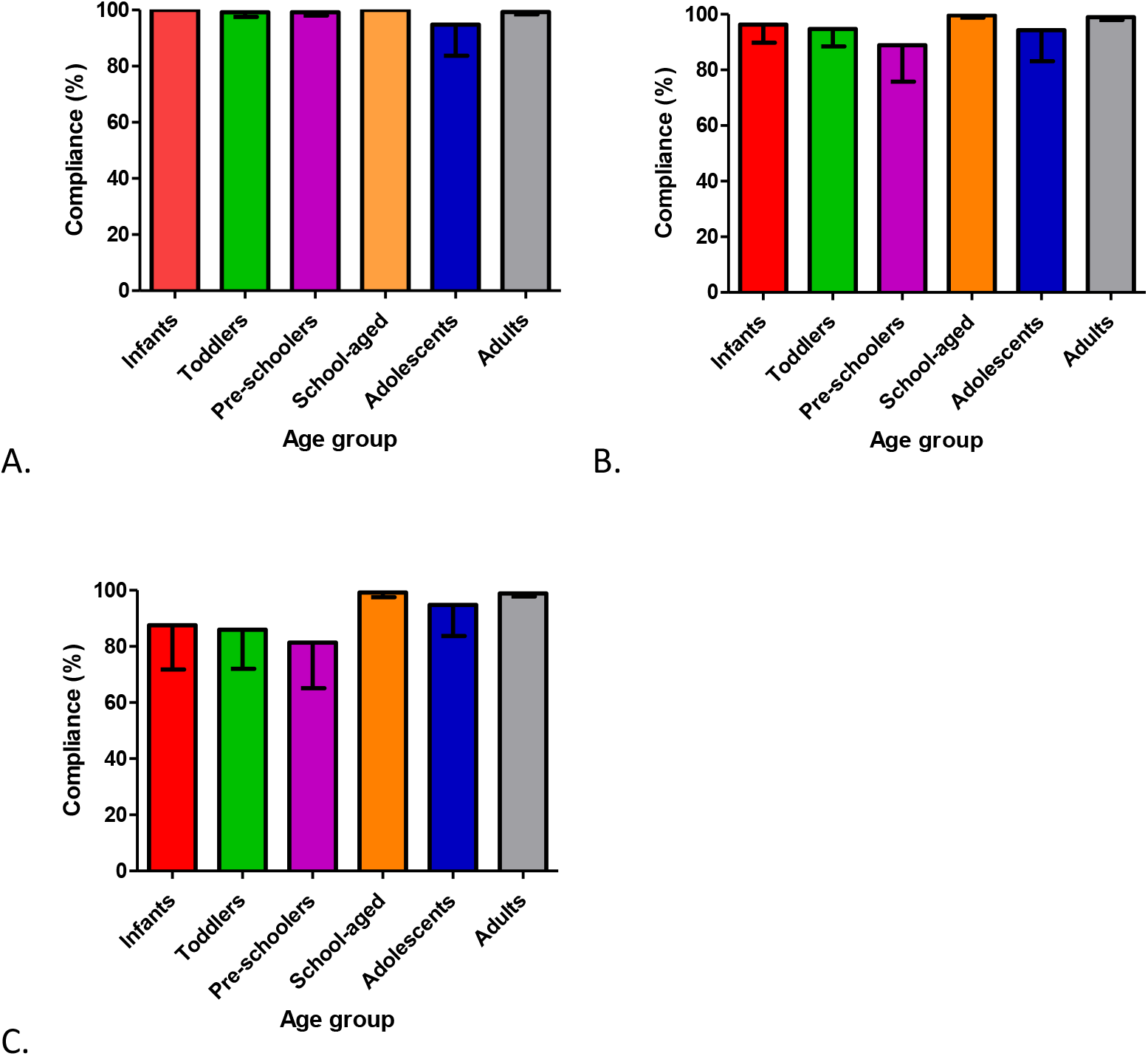
Participant compliance with collection of (A) hand swab, (B) saliva and (C) nasal lining fluid samples at home. Data is expressed as percentage of the mean of the compliance of each participant per age group, with compliance the number of samples collected from a participant as a proportion of the number of samples expected based on the number of weeks their family participated in the study. Bars represent 95% Confidence Intervals. Infants n=9, toddlers n=11, pre-schoolers n=21, school-aged n=21, adolescents n=15, adults n=80.

### Family and participant acceptability to methodology

Only families who completed all six months of sampling and returned their acceptability questionnaire were included in this analysis (n=33). Twenty-five families (76% CI [58%,89%]) indicated their overall acceptance to our methodology; families who did not find it acceptable were more likely to have at least one child <5 years (Figure 4A). Overall, 73% CI [64%,80%] of individual participants accepted our methodology. Participant acceptability varied with age, with acceptability highest amongst adults (86% CI [76%,94%]) and lowest for toddlers (30% CI [7%,65%]) (Figure 4B). Our methodology was found to be fast to complete (76% CI [55%,91%]), easy for sample collection (76% CI [55%,91%]) and painless (60% CI [39%,79%]). Furthermore, 20% CI [7%,41%] of families commented that sampling at home was preferable to traveling to a testing center. Based on the most important reason (with a score of 1), acceptability varied with family composition with fast completion (29% CI [8%,58%]), ease of sample completion (43% CI [18%,71%]), painless collection (21% CI [5%,51%]) and sampling at home (7% CI [0.2%,34%]) being the primary reasons for accepting our methodology (Figure 4C).

**Figure 4.**
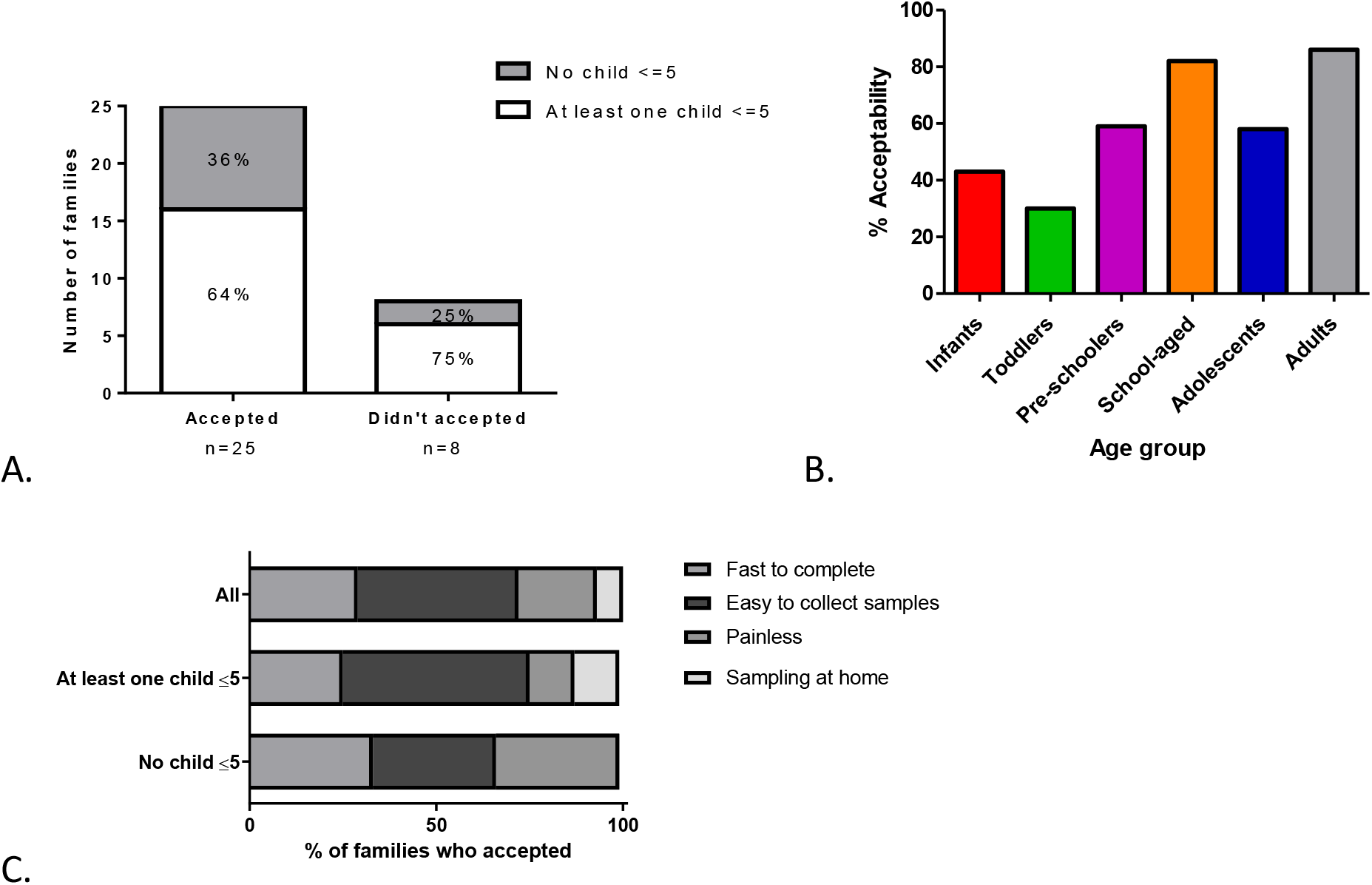
Acceptability of the methodology expressed by (A) number of families, (B) % participants per age group. (Infants n=7, toddlers n=10, pre-schoolers n=17, school-aged n=17, adolescents n=12, adults n=66) **and (C) primary reasons for acceptance per family composition**.

### Participant acceptability to sample type

Hand swabs were universally accepted, and most participants also accepted saliva sampling (Figure 5A). Nasal sampling was the most challenging, with a steady decline in acceptability with decreasing age and only 14% CI [0.4%,58%] acceptability in the youngest age group of infants (Figure 5B).

**Figure 5.**
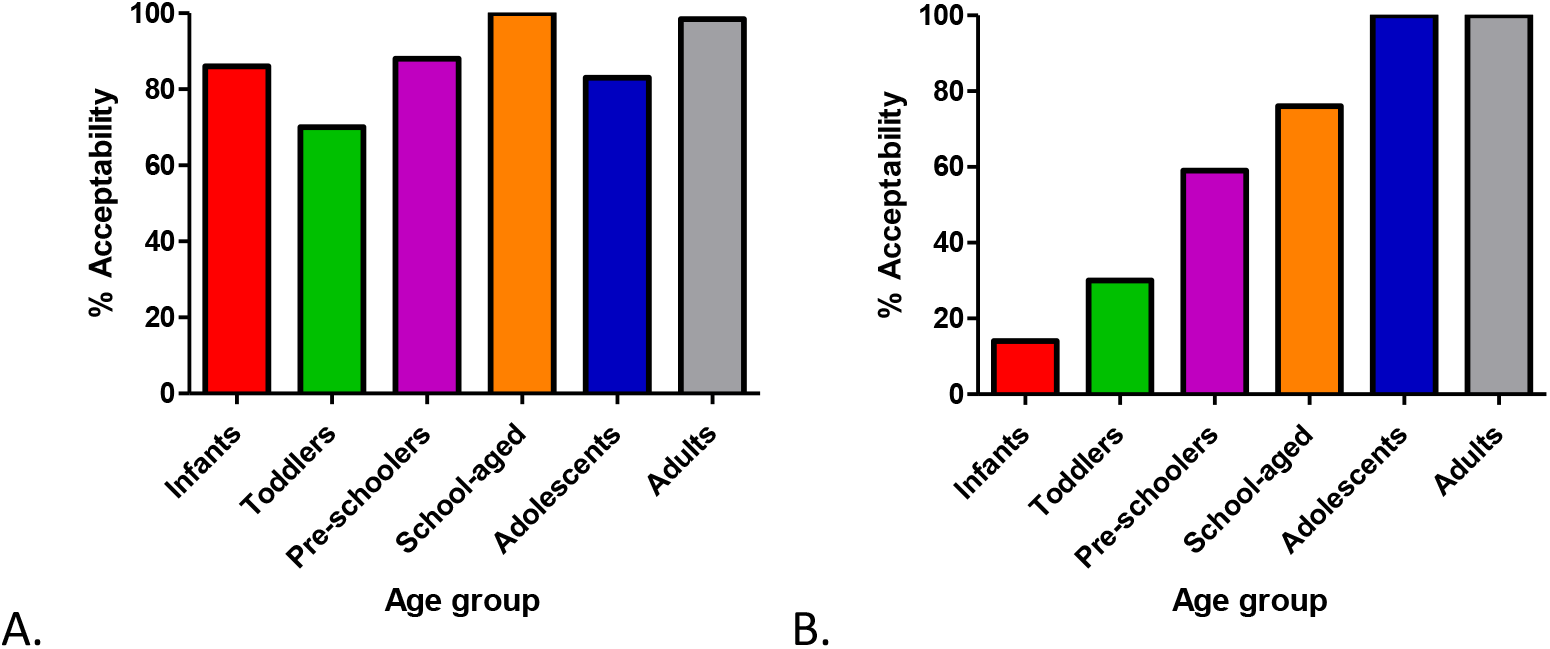
Participant acceptability to (A) saliva and (B) nasal lining fluid sampling per age group. Data is expressed as percentage of the number of participants accepted in one age group divided by the total number of participants in that age group. Infants n=7, toddlers n=10, pre-schoolers n=17, school-aged n=17, adolescents n=12, adults n=66.

## Discussion

We have developed a methodology comprising of minimally invasive biological self-sampling with questionnaires for monitoring of the human oral, nasal and hand microbiota, and assessed its acceptability in a family setting after six months. We demonstrated that our methodology was acceptable for many parents and children across a wide age range, with highest acceptability amongst adults and school-aged children. To our knowledge this is the first study that employs longitudinal, minimally invasive sampling at home in a family setting, with collection of samples from three different human niches.

The approach of sample self-collection is not new. Self-taken URT swabs have previously been used for diagnostic purposes, especially during the COVID-19 pandemic where their use increased the testing rate, preserved scarce personal protective equipment (PPE), and reduced illness in health care workers [25]. Recent research has demonstrated that nasal swab collection by trained staff is not superior to self-collected or parent-assisted swabs [26]. Importantly, the laboratory yield of samples was not determined by who took the sample but by the anatomical site from when the sample was taken. Self-collection of saliva has been also used for diagnosing SARS-CoV-2 and is considered by some as the best method [27]. This method does not cause discomfort or pain, can reduce healthcare personnel exposure by avoiding coughing, sneezing, and/or aerosolization during sampling and requires fewer consumables, offering a significant benefit during supply shortages.

In our study, sampling at home was one of the reasons that participants accepted our methodology indicating the usefulness of our approach compared to clinical visits. As expected, both saliva and hand sampling were acceptable to participants of all ages, whereas nasal sampling was accepted mostly by adults and older children (>5 years). Nasal and nasopharyngeal swab collection are associated with a certain amount of pain and discomfort, making them especially unpopular with young children. Even though SAM is less invasive than a swab, infants and toddlers were distressed during nasal sampling procedure as reported by their parents [24]. Therefore, their acceptability was low for these age groups, with a range of time taken for SAM sampling between 1 second and 2 minutes [24]. However, compliance was high (above 80%) as parents kept trying to collect samples. A study using SAM in hospitalized children found that 30 seconds of sampling yielded good results [18]. Limiting the time taken for sampling to 30 seconds may improve acceptability in young children, though it is likely that there some will not tolerate sampling even for this shorter period or who refuse sampling over time.

Our study has several strengths. It was conducted during and after strict COVID-19 restrictions thus we capture the feasibility of our methodology in the challenging context of a pandemic. We recruited 77 children between 2 months and 17 years of age, with a good distribution across the ages. While the number of children in each age group was limited, the numbers were sufficient to observe substantial differences in acceptability and compliance between the groups.

A major limitation of our work is that participating adults had much higher annual household incomes and educational status than the national average despite Liverpool being one of the most deprived local authorities in England. Socioeconomic status has previously been seen to affect participation in a study of one-off self-swabbing [28]. Additionally, a large proportion of the adults in our study worked in medical, research or life science professions and may be more likely to find our methods acceptable. Because personal contact was the most successful recruitment method, it is also possible that many participants were more likely to comply with the methodology because of the personal connection. Therefore, our findings may not be generalisable to the UK population.

At-home specimen collection for respiratory pathogens enables community sampling and households have been highlighted as an important source of microbial spread within the community [29-31]. Thus, considerations of family members of different ages on comfort and ease with which samples can be obtained are critical for informing future large surveillance studies of microbial prevalence and transmission. Overall, the sampling methods used had a high level of acceptance amongst families in this study. Further research is needed to assess acceptability in populations with lower socioeconomic status and to adapt sampling methods for nasal fluid in younger children.

## Supporting information

Suppl Doc 1

Suppl Doc 2

Suppl Doc 3

Suppl Doc 4

## Data Availability

All data produced in the present study are available upon reasonable request to the authors

## Acknowledgments

The authors would like to thank all participants enrolled in the study for their time and effort especially during the pandemic. We would also like to thank Alder Hey Children’s Hospital for their collaboration as a study recruitment site, as well as Kelly Davies and Kelly Convey for their invaluable assistance in trial set-up and management. The study was supported by the Director’s Catalyst Fund 2020 awarded to Dr Nikolaou and the UKRI Strength in Places Fund awarded to Prof Ferreira.

