## Supplementary material for "Family Research of Microbes Linked to Respiratory Infections (FAMILY Micro) observational study: Assessing the use of minimally invasive self-sampling methodologies at home for long-term monitoring of the oral, nasal and hand microbiota of adults and children within UK families": Suppl Doc 1

**Sample collection kit contents**

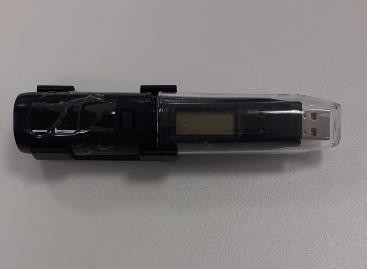

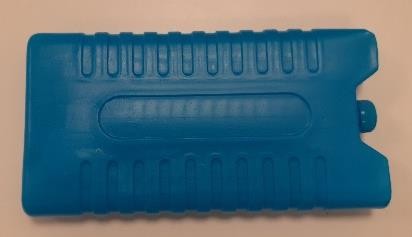

#### Thermometer Cool pack

ALWAYS keep in FREEZER with samples	FREEZE before use

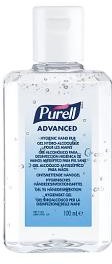

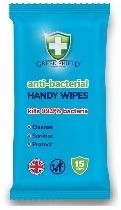

#### Hand sanitizing gel Disinfectant wipes

**HAND SWAB KIT	NASAL STRIP KIT**

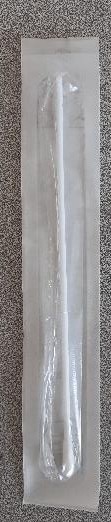

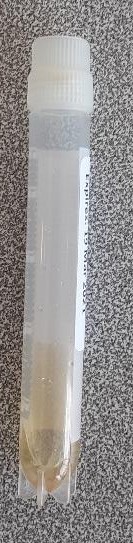

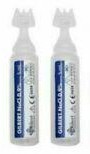

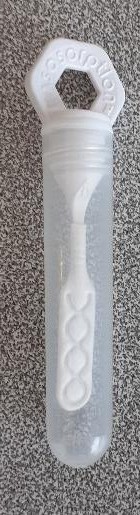

#### Swab Storage tube Saline Nasal strip with container

**KEEP IN FRIDGE**

**SALIVA KIT**

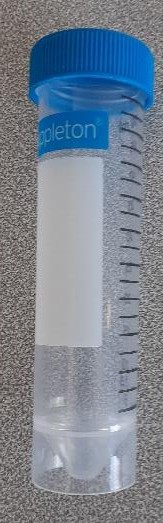

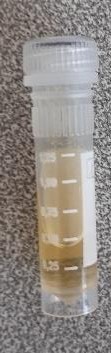
**Adults and children >6 years**

#### Saliva tube Preservative liquid tube

**KEEP IN FRIDGE**

#### Infants and Toddlers

a) **Swab storage tube (KEEP IN FRIDGE)**

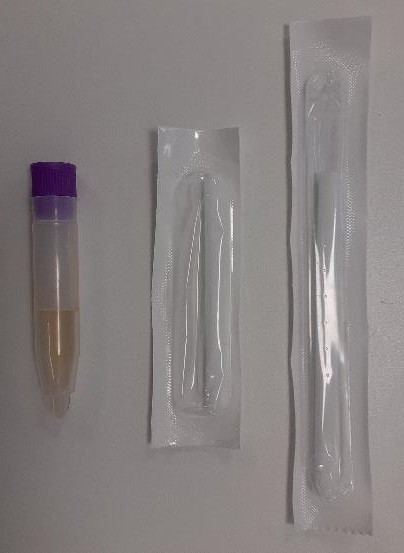

***a)***

***b)***

***c)***

#### b) Infant’s swab (28 days – 1 year)

c) **Children’s swab >1 year**

### ATTENTION: KEEP ALL DEVICES OUT OF THE REACH OF CHILDREN AND SUPERVISE AT ALL TIMES DURING SAMPLE COLLECTION!!
