## Supplementary material for "Family Research of Microbes Linked to Respiratory Infections (FAMILY Micro) observational study: Assessing the use of minimally invasive self-sampling methodologies at home for long-term monitoring of the oral, nasal and hand microbiota of adults and children within UK families": Suppl Doc 2

#### Dear Participant

**Information sheet for home collection of samples in the *FAMILY MICRO* research study.**

We are studying the change in type and numbers of bacteria in the nose and mouth of family members over time and exploring which bacteria can be transferred via our hands. We will use this information to better prevent and treat respiratory infections in the future. We are asking you to collect saliva, nasal samples, and hand swabs at home every 2 weeks for 6 months, on the days listed on the schedule below.

It is very important that the samples are taken from all family members on the same day of the study week and a maximum of one day before or after the day indicated on the schedule planner. The time of collection does not matter as long as all samples are collected on the same day. Time will be recorded at the “Home Sampling Practicality Questionnaire” ONLY to assess practicality of the method used (especially for children). If you are unavoidably delayed (or one of the family members cannot collect samples on the same day) please still collect the sample and write the exact day taken in the “Schedule Planner”. We ask you to take a photo with your phone of all samples just after all family members have collected their samples and send it to us via WhatsApp.

#### General Instructions:

1) Set a weekly alarm on your phone so you will not forget to take the samples on that day.
2) Each tube is marked with the study week that the sample is due (Week0-Week22, planned on the form below). Each member of the family has their own tubes labelled as PARENT A (purple), PARENT B (green), CHILD A (yellow), CHILD B (orange). EACH MEMBER HAS TO USE THEIR OWN TUBES THROUGHOUT THE STUDY.
3) When you have collected each sample, they must be stored in your freezer in the Tupperware box provided. If for any reason, a family member is not at home when they need to take the sample, please use the cool pack in the cool bag or place samples in a fridge if available. Once back home, place the samples in your freezer.
4) Store the cool pack in the freezer when you are at home if needed in the following days.
5) Complete the “Schedule Planner” and “Home Sampling Practicality Questionnaire”

each time you collect a sample.

6) For infants and children who cannot spit please use the paediatric swabs for saliva collection.

### VERY IMPORTANT

**After taking all samples on the same day, REMEMBER to:**

1. Photograph the samples using your phone immediately after all members have taken them. If samples have not been taken at the same time, please send pictures after the samples have been collected and before freezing. Forward the photo to TEL: 07748417604 then store the samples in the freezer.
2. Fill in your “Schedule Planner” and “Home Sampling Practicality Questionnaire”. Keep all samples and accompanying forms to be returned to us when required (half of the samples at 3 months and the other half at the end of the study).

### Thank you for taking part in this study!

**THE DAY OF SAMPLE COLLECTION**

Please follow these steps if taking samples at the same time:

1. Take your hand swab sample.
2. Use a disinfectant wipe to clean the area that you will collect samples.
3. Wash your hands for 20 seconds with soap and warm water.
4. Help your children take their hand swab samples (if help is needed). If you do this step, then sanitize your hands before proceeding to step 5.
5. Take your nasal strip sample.
6. Sanitize your hands.
7. Help your children take their nasosorption swab samples (if help is needed). If you do this step, then sanitize your hands before proceeding to step 8.
8. Take your saliva sample.
9. Sanitize your hands.
10. Help your children take their saliva samples. IMPORTANT: Make sure you have sanitized your hands before helping infants, toddlers, and pre-schoolers to take their saliva sample (before touching the paediatric swab).
11. At the end, wash your hands for 20 seconds with soap & warm water.

NB: You can take your samples at anytime on the same day of the study week if you do not wish to take them altogether. REMEMBER to wash your hands at the beginning before taking samples and to sanitize your hands in between the different samples collected or individuals (e.g. between you and your children).

**WASH YOUR HANDS:**

- Wet your hands with clean, running warm water and apply soap.
- Lather your hands by rubbing them together with the soap. Lather the backs of your hands, between your fingers, under your nails and the tops of your hands. Scrub your hands for at least 20 seconds.
- Rinse your hands well under clean, running water for 10 seconds.
- Dry your hands using a clean towel.
- Turn off tap using the towel.

**SANITIZE YOUR HANDS:**

- Apply a small amount of the instant hand sanitising gel to the palm of one dry hand.
- Rub your hands together.
- Rub the gel over all the surfaces of your hands and fingers until your hands are dry. This should take around 20 seconds.
