## Supplementary material for "Family Research of Microbes Linked to Respiratory Infections (FAMILY Micro) observational study: Assessing the use of minimally invasive self-sampling methodologies at home for long-term monitoring of the oral, nasal and hand microbiota of adults and children within UK families": Suppl Doc 3

**SCHEDULE PLANNER	Date study commenced: Date Study completed:**

| **Study**  **week** | **Planned date** | **Actual date taken**  (if different from the planned date) | **Hand Swab sample** | | | | | **Nasal Strip sample** | | | | | **Saliva spit/swab sample** | | | | | **Photograph sent, store samples in freezer, questionnaire filled in (tick)** |
| --- | --- | --- | --- | --- | --- | --- | --- | --- | --- | --- | --- | --- | --- | --- | --- | --- | --- | --- |
|  |  |  | **Parent A** | **Parent B** | **Child A** | **Child B** | **Child C** | **Parent A** | **Parent B** | **Child A** | **Child B** | **Child C** | **Parent A** | **Parent B** | **Child A** | **Child B** | **Child C** |  |
| **Week 0** |  |  |  |  |  |  |  |  |  |  |  |  |  |  |  |  |  |  |
| **Week 2** |  |  |  |  |  |  |  |  |  |  |  |  |  |  |  |  |  |  |
| **Week 4** |  |  |  |  |  |  |  |  |  |  |  |  |  |  |  |  |  |  |
| **Week 6** |  |  |  |  |  |  |  |  |  |  |  |  |  |  |  |  |  |  |
| **Week 8** |  |  |  |  |  |  |  |  |  |  |  |  |  |  |  |  |  |  |
| **Week 10** |  |  |  |  |  |  |  |  |  |  |  |  |  |  |  |  |  |  |
| **Week 12** |  |  |  |  |  |  |  |  |  |  |  |  |  |  |  |  |  |  |
| **Week 14** |  |  |  |  |  |  |  |  |  |  |  |  |  |  |  |  |  |  |
| **Week 16** |  |  |  |  |  |  |  |  |  |  |  |  |  |  |  |  |  |  |
| **Week 18** |  |  |  |  |  |  |  |  |  |  |  |  |  |  |  |  |  |  |
| **Week 20** |  |  |  |  |  |  |  |  |  |  |  |  |  |  |  |  |  |  |
| **Week 22** |  |  |  |  |  |  |  |  |  |  |  |  |  |  |  |  |  |  |
