## Supplementary material for "Family Research of Microbes Linked to Respiratory Infections (FAMILY Micro) observational study: Assessing the use of minimally invasive self-sampling methodologies at home for long-term monitoring of the oral, nasal and hand microbiota of adults and children within UK families": Suppl Doc 4

**Home Sampling Acceptability Questionnaire**

**TO BE COMPLETED AT THE END OF THE STUDY**

Instructions: Please read the questions carefully and select the right box(es). There is no right or wrong answer. Just indicate by marking the appropriate box what you have experienced and felt.

1. Do you find home sampling acceptable as a method for collecting samples from families (tick one box only *🗹* )?

□Yes (please go to question 2)	□ No (please go to question 3)

2. If you answered YES to Question 1, please indicate your top three reasons for accepting home sampling by putting a number in the box: 1 = most important feature, 2 = next most important feature, 3 = subsequent most important feature.

□Fast to complete	□Easy to collect samples

□Does not cause pain	□Does not cause discomfort

□Easily applicable to children (specify the age………………………………………………………………………….)

□Other, specify………………………………………………………………………………………………………………………….

3. If you answered NO to Question 1, please indicate your top three reasons for NOT accepting home sampling by putting a number in the box: 1 = most important feature, 2 = next most important feature, 3 = subsequent most important feature.

□Takes time to complete	□Not easy to collect samples

□Causes pain	□Causes discomfort	□Not easily applicable to children (specify the age……………………………………………………………………)

□Other, specify………………………………………………………………………………………………………………………….

4. The answers to questions 1-3 agreed by all family members.

□Yes	□No. Specify who does not agree (e.g. Parent B) and why …………………………………………………………………………………………………………………………………………………………………………………………………………………………………………………………………………………………………………………………
