## Supplementary material for "Family Research of Microbes Linked to Respiratory Infections (FAMILY Micro) observational study: Assessing the use of minimally invasive self-sampling methodologies at home for long-term monitoring of the oral, nasal and hand microbiota of adults and children within UK families": Suppl Table 1

**Supplementary Tables**

**Table S1. Inclusion and exclusion criteria.**

| ***Inclusion Criteria*** | |
| --- | --- |
| ***Age*** | Adults 18-60 years and 1-3 children 28 days - 17 years |
| ***English language*** | Adults with fluent spoken English to ensure a comprehensive understanding of the research project and their proposed involvement |
| ***Consent*** | Adult capacity to give informed consent |
| ***Study’s requirements*** | All family members, both parents and children, must agree to participate in the study and live in the same household during the study period. Drop out of a member during the study was acceptable. |
| ***Exclusion Criteria*** | |
| ***Research*** | Currently involved in a CTIMP or any trial that can affect the microbiome, at the discretion of the study Dr |
| ***Health History*** | Daily medications that may affect the microbiome e.g. long-term antibiotics or immunosuppressants including steroids |
|  | Recent immunosuppressants or antibiotics within the last 28 days. In this case, recruitment was delayed |
|  | History of respiratory infections requiring hospitalisation |
|  | Current severe acute respiratory infection |
|  | Disease or syndrome associated with altered immunity or altered respiratory or gut microbiome (Chron’s, ulcerative colitis or diabetes, asthma with regular medication/COPD) or other RTI conditions (including cystic fibrosis or bronchiectasis) |
|  | Children with cognitive disabilities that lead to an inability to comply with study sampling |
| ***Pregnancy*** | Pregnant at recruitment, however pregnancy was accepted during the study |
